# Time-Varying Risk of Death After SARS-CoV-2-Infection in Long-Term Care Facility Residents: A Matched Cohort Study

**DOI:** 10.1101/2022.03.10.22272097

**Authors:** Marcel Ballin, John P.A. Ioannidis, Jonathan Bergman, Miia Kivipelto, Anna Nordström, Peter Nordström

## Abstract

**Background:** SARS-CoV-2 confers high risk of short-term death in residents of long-term care (LTC) facilities, but longer-term risk among survivors is unclear.

**Methods:** We extended the follow-up period of a previous, propensity score-matched retrospective cohort study based on the Swedish Senior Alert register. N=3731 LTC residents with documented SARS-CoV-2 until 15 September 2020 were matched to 3731 uninfected controls using time-dependent propensity scores on age, sex, health status, comorbidities, and prescription medications. In a sensitivity analysis, matching included also geographical region and Senior Alert registration time. The outcome was all-cause mortality over 8 months (until October 24, 2020). The absolute risk of death was examined using Kaplan-Meier plots. Hazard ratios (HR) for death over time were estimated using flexible parametric models with restricted cubic splines. Cox regression was used to estimate HRs and 95% confidence intervals (CIs) in 30-day intervals of follow-up until 210 days.

**Results:** The median age was 87 years and 65% were women. Excess mortality was highest 5 days after documented infection (HR 19.1, 95% CI, 14.6-24.8), after which excess mortality decreased. From the second month onwards, mortality rate became lower in infected residents than controls. The HR for death during days 61-210 of follow-up was 0.41 in the main analysis (95% CI, 0.34-0.50) and 0.76 (95% CI, 0.62-0.93) in the sensitivity analysis. Median survival of uninfected controls was 1.6 years, which was much lower than the national life expectancy in Sweden at age 87 (5.05 years in men, 6.07 years in women).

**Conclusions:** No excess mortality was observed in LTC residents who survived the acute SARS-CoV-2 infection. Life expectancy of uninfected residents was much lower than that of the general population of the same age and sex. This suggests that LTC resident status should be accounted for in years-of-life-lost estimates for COVID-19 burden of disease calculations.

**Impact statement:** We certify that this work is novel. This research adds to the literature by showing there was no excess mortality observed in long-term care facility residents who survived the acute SARS-CoV-2 infection, and that life expectancy of uninfected residents was much lower than that of the general population of same age and sex. This has major repercussions for estimation of years of life lost in infected long term care facility residents.

**Key points:**

- SARS-CoV-2 infection sharply increased mortality risk among residents of long-term care (LTC) facilities in the first month.
- After the first month, the mortality risk in infected residents rapidly returned to baseline and dropped below the mortality risk of uninfected controls, where it remained lower for 8 months of follow-up.
- Median survival of uninfected controls was 1.6 years, which was much lower than national life expectancy in Sweden at age 87.

**Why does this matter?:**

- Whereas LTC residents who recover from SARS-CoV-2 infection may be concerned about having residual debilitation caused by the infection, we found no excess mortality was in those who survived the acute infection.
- Because life expectancy of uninfected residents was much lower than that of the general population of same age and sex, LTC resident status should be accounted for in estimations of years of life lost.

## Introduction

During the coronavirus disease 2019 (COVID-19) pandemic, many of the COVID-19 deaths that occurred in high-income countries were seen in long-term care (LTC) facilities,^1^ where case fatality rates were 10-40% or even higher.^2,3^ We have previously reported that 30-day mortality in Swedish LTC was 40% in residents infected with severe acute respiratory syndrome coronavirus 2 (SARS-CoV-2) versus 6% in matched, non-infected controls in the first wave of the pandemic.^4^ A natural follow-up question to ask is whether SARS-CoV-2 also increases the risk of death beyond the acute period of 30 days, i.e. whether it has long-term effects on mortality in LTC residents who recover from infection. A major concern is that LTC residents who recover from SARS-CoV-2 infection may have residual debilitation caused by the infection. If so, this may affect also their life expectancy beyond the acute phase of the infection. Moreover, it would be interesting to estimate the loss of life expectancy in LTC residents infected with SARS-CoV-2. We set out to answer these questions by extending the follow-up period in our previous analysis from 30 days to 8 months.

## Methods

### Study design and population

The present study offers extended follow-up on a propensity score-matched retrospective cohort study.^4^ The basic study design and of the selection of exposed (infected) and unexposed (uninfected control) residents has been described in detail previously in the publication presenting 30-days of follow-up.^4^ In brief, data on Swedish LTC residents were obtained from Senior Alert, a database of health assessments performed in older adults aged ≥65 years.^5^ All residents of LTC facilities in Sweden registered in Senior Alert were eligible to be considered. Senior Alert collects health data on various conditions in adults aged ≥65 years. Senior Alert captures an estimated 73% of all Swedish LTC facility residents. This study was approved by the Swedish Ethical Review Authority (no. 2020-02552), which waived the informed consent requirement given the retrospective nature of the study. The study was performed in accordance with the Declaration of Helsinki.

### Cohort construction and matching

We selected LTC residents who had a record in Senior Alert from 2019 or 2020; the latest record during these years was used, whenever there were multiple records. Among these, we identified 3731 LTC residents with a documented SARS-CoV-2 infection until September 15, 2020. We excluded SARS-CoV-2 infected residents that did not have a record in Senior Alert within a year prior to date of testing or confirmed infection (whichever came first or was available) and those where dates of testing and confirmed infection were both unavailable. Data on SARS-CoV-2 infections were obtained from the national SmiNet registry to which reporting is mandatory according to Swedish law.

Each infected resident was matched 1:1 to a control resident on age, sex, body mass index, health status, comorbidity, and prescription medication use, using time-dependent propensity scores. This enables matching when the exposure (date of documented SARS-CoV-2 infection) do not coincide with the time of cohort entry (date of Senior Alert record). With time starting at the date of the Senior Alert record, a Cox model calculated a propensity score for the propensity to contract SARS-CoV-2 based on all variables shown in Supplemental Table 1. Each infected resident was matched to the control with the closest propensity score among those who were still alive when the SARS-CoV-2 case occurred (counting time since the Senior Alert date). Matching was done sequentially, starting with the first case (smaller number of days since cohort entry) and proceeding with cases with increasingly larger number of days since cohort entry. Diagnoses and medications were used as time-varying covariates in the Cox regression model. Information on comorbidities was obtained from the Swedish National Patient Register and for cancer from the Swedish Cancer Register. Information on recent use of medications (prescriptions in 2019-2020) came from Senior Alert and the Swedish Prescribed Drug Register. In addition, in order to conduct a sensitivity analysis to examine whether the results were confounded by secular trends and geographic variations in mortality, we performed an additional matching which included also the geographical region (21 categories) and the date (year and month) of Senior Alert registration. Data linkage across registers were performed using pseudo-anonymised Personal Identification Numbers.

### Outcome

The study outcome was all-cause mortality (until October 24, 2020). These data were obtained from the national Swedish Cause of Death Register. While in our previous study we only assessed 30-day mortality,^4^, the extension to 8 months in the present study allows to get a more complete picture of the mortality risk of this frail population, while at the same time it largely excludes the subsequent waves and also the COVID-19 vaccination period which may have further affected mortality risk in this population.

### Statistical analysis

In both the main and sensitivity analyses for death risk, all-cause mortality was considered as the outcome of interest and the starting date for follow-up was the SARS-CoV-2 documentation date in cases and the corresponding date (in days since cohort entry) in controls. Follow-up time in days was calculated as censor date (24 October 2020 or death whichever came first) minus baseline date + 1 day. This was done so that the baseline date could also be included in the follow-up time and analysis (thus, a person would be able to die on the same date as they were documented to be infected). The absolute risk of death was examined using Kaplan-Meier plots. The hazard ratio (HR) for death was plotted over time using flexible parametric models with restricted cubic splines (4 knots in default positions). HRs and 95% confidence intervals (CIs) were also estimated using Cox regression for 30-day intervals of follow-up until 210 days. To adjust for matching, we calculated 95% CIs in the Cox models and the flexible parametric models using robust standard errors. All analyses were performed using Stata MP version 16.1 for Mac (StataCorp, College Station, TX).

## Results

### Baseline characteristics

Baseline characteristics are shown in Supplemental Table 1. The median age was 87 years, 65% were women, and comorbidities were common. In the main analysis, median (interquartile range [IQR] baseline date for infected residents was 27 Apr 2020 (10 Apr to 22 May), median (maximum) follow-up was 129 (246) days and there were 1713 deaths. For controls, median (IQR) baseline date was 12 Apr 2020 (16 Dec 2019 to 30 Jun 2020), median (maximum) follow-up was 146 (641) days and there were 899 deaths. In the sensitivity analysis, for infected residents, median (IQR) baseline date was 26 Apr 2020 (IQR 10 Apr to 21 May), median (maximum) follow-up was 130 (246) days and there were 1640 deaths. For controls, median (IQR) baseline date was 28 Apr 2020 (9 Apr to 23 May), median (maximum) follow-up was 173 (249) days and there were 536 deaths. The median age was 87 years, 65% were women, and comorbidities were common (Supplemental Table 1).

### Mortality analyses

As previously reported, SARS-CoV-2 was associated with a sharp, early increased risk of death: 40% versus 6% within 30 days (1487/3731 versus 211/3731). However, extending the follow-up period showed that the risk soon plateaued (Figure 1A). Similar results were seen in the sensitivity analysis (Figure 1B).

**Figure 1.**
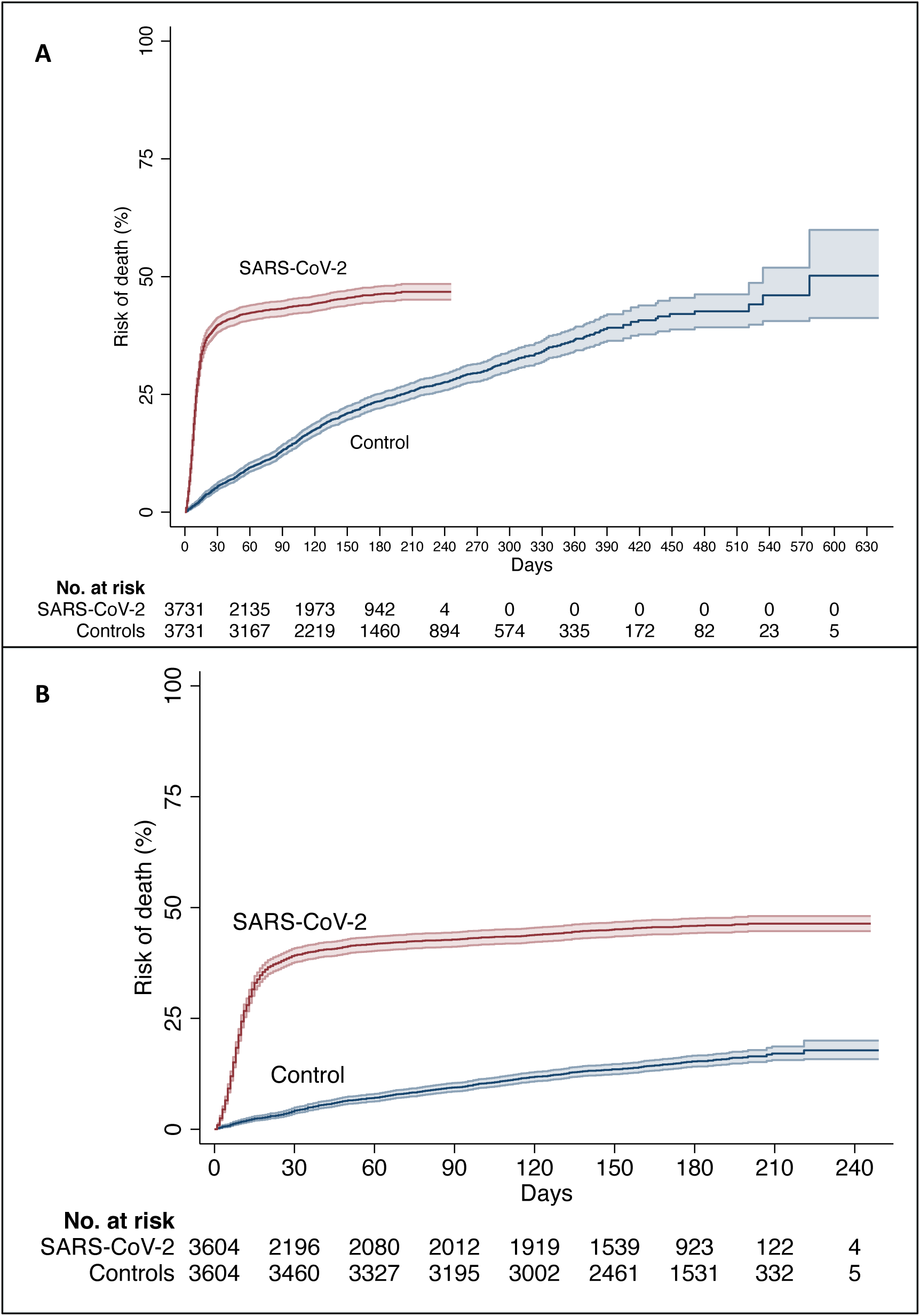
Risk of death in residents with SARS-CoV-2 and controls in the main analysis (A) and in the sensitivity analysis (B). The coloured areas show the 95% confidence interval.

Survival of controls at 210 days was 74.3% (72.6%-75.9%) in the main analysis and 82.9% (81.3%-84.4%) in the sensitivity analysis. Median survival of controls was 577 days. Median survival was also 577 days among the controls who were matched to the 1487 infected residents who died in the first month. Survival of these 1487 controls was similar to the survival of the remaining 2242 controls, for example their survival at 210 days was 72.5% versus 75.4%.

In the main analysis, peak HR (19.1 (95% CI, 14.6-24.8)) occurred at 5 days after documented infection. HR was high in the first month, decreased below 1.0 early in the second month, and remained below 1.0 for the remaining duration of follow-up (Figure 2a). In the sensitivity analysis, peak HR was 21.5 (95% CI, 15.9-29.2). Again, HR decreased sharply but took a bit longer to drop below 1.0 (after the second month) and remained below 1.0 afterwards (Figure 2b).

**Figure 2.**
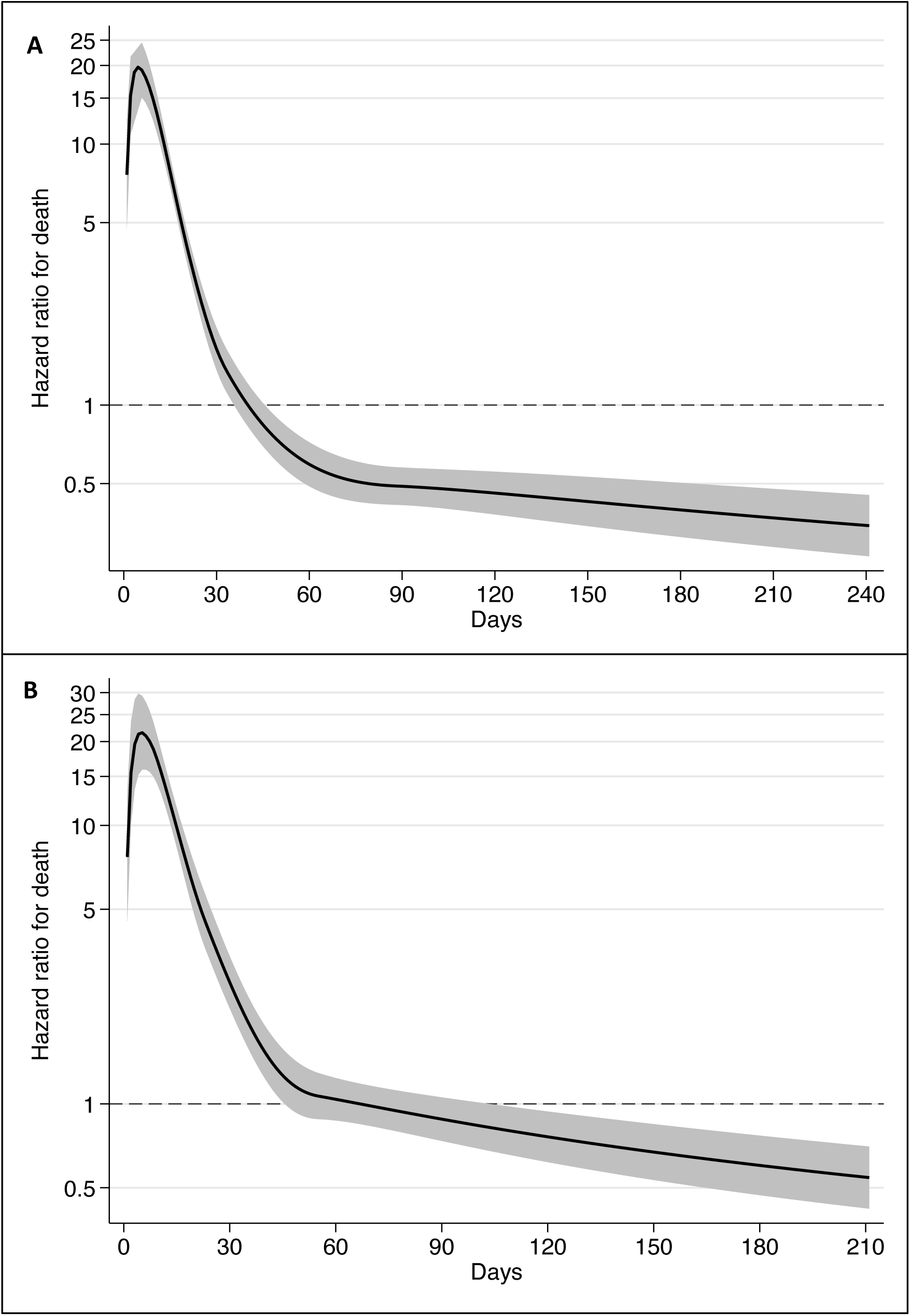
Hazard ratio for death in residents with SARS-CoV-2, as compared with controls in (A) the main analysis and (B) the sensitivity analysis. The coloured areas show the 95% confidence interval

In the main analysis, for 0-30 days, there were 1487 deaths among infected residents (17.57 deaths per 1000 person-days) versus 211 in uninfected controls (1.88 deaths per 1000 person-days), resulting in a HR of 8.81 (7.64-10.15). For 31-60 days, there were 93 deaths (1.42 per 1000 person-days) versus 144 (1.42 per 1000 person-days) (HR 1.00 (0.77-1.30)). For 61-90 days the respective numbers were 33 (0.55) versus 121 (1.38) (HR 0.38 (0.26-0.55)). For 91-120 days, the respective numbers were 38 (0.63) versus 126 (1.72) (HR 0.36 (0.25-0.52)). A similar pattern was seen for 121-150 days (HR, 0.52 (0.36-0.76)), 151-180 days (HR 0.47 (0.28-0.79)), and 181-210 days (HR 0.29 (0.10-0.83)).

During the 61-210 days follow-up, there were 133 deaths among infected residents (0.58 per 1000 person-days) versus 420 deaths among the uninfected controls (1.37 per 1000 person-days), with the HR being 0.41 (0.34-0.50). In the sensitivity analysis, during 61-210 days of follow-up, there were 131 deaths (0.59 per 1000 person-days) in infected versus 278 deaths (0.78 per 1000 person-days) in controls, with the HR being 0.76, 95% CI, 0.62-0.93).

## Discussion

In this extended follow-up analysis of mortality in SARS-CoV-2-infected versus uninfected control LTC residents, we found that mortality risk peaked during the first week of documented infection, after which it rapidly decreased. Mortality remained elevated for the first month after infection, but then reverted back to baseline levels (i.e., control levels) before it dropped below baseline levels, where it remained at low levels for the remaining duration of follow-up (up to 8 months). These results suggest that SARS-CoV-2 does not reduce the life expectancy of LTC residents who survive the acute period of the disease. Despite concerns that infected residents who survive may have persistent residual debilitation that might enhance their subsequent death risk, we saw the opposite: death risk decreased in longer-term follow-up. This suggests that deaths due to COVID-19 in LTC facilities in Sweden during the first wave probably resulted in average loss of life expectancy of less than 1.6 years on average. This figure is much lower than the life expectancy in the general Swedish population, which in 2019 was 5.05 years for men and 6.07 years for women at the age of 87 (the median age in our study).^7^

Calculations of burden of disease due to COVID-19 often use age- and sex-adjusted life expectancies to calculate years-of-life-lost; however, without properly accounting for LTC residence and general health. Our findings suggest that such an approach can yield massively inflated estimates.^8^ Adjustment for comorbidities has been shown to decrease the number of years-of-life-lost in some studies.^9-11^ However, the change is typically modest (e.g. in the range of 1 years) and much smaller than what we observed in the LTC resident population that we evaluated. It is possible that in most studies, information on comorbidities is not available in sufficient granularity and accuracy regarding severity. E.g. “kidney disease” would carry very different risk connotations depending on the stage and severity. LTC resident status is a surrogate for increased frequency and severity of many comorbidities and of overall frailty. Therefore, it should be taken into account as a first correction for any years-of-life-lost estimates for COVID-19 burden of disease calculations.

Calculations accounting for LTC status and also properly adjusting for comorbidities and their severity may also lead to much lower estimates than some other increasingly used approaches such as the Global Burden of Disease Reference Life Table^12^ - also known as Theoretical Minimum Risk Life Table. This life table is an “aspirational” construct: it assumes an idealized situation with very low risk of death. According to this table, life expectancy is 88.9 years at birth, 9.99 years at age 85, 5.92 years at age 90, and 5.92 years at age 95.^13^ Using this popular aspirational life table would probably overestimate by 5-10-fold the years-of-life-lost for SARS-CoV-2-deceased residents in LTC facilities. Aspirational life tables have been promoted as a way to standardize burden of disease calculations across different countries. However, in the case of diseases like COVID-19 they could lead to grossly misleading inferences.

Estimates of survival in residents of LTC facilities preceding the COVID-19 pandemic also agree with very limited median survival of nursing home residents, e.g. 541 days in one study in the United Kingdom^14^ and 2 years in a study of residential care entrants in New Zealand.^15^ In Sweden, previously published data^16^ on the survival of elderly people who moved into institutionalized care in an area of Stockholm (N=1103) suggested that, on average, the median survival after moving to institutionalized care declined between 2006 and 2012 from 764 to 595 days. For the lower percentiles, the decrease was very large, e.g. for the 30th percentile, the length of stay declined from 335 days in 2006 to 119 days in 2012, and in 2012 10% died within just 8 days. A widening survival gap (due to shortening survival in nursing home residents) versus community-dwelling elderly has also been documented in a 10-year study in England.^17^ Another study^18^ evaluated all deaths in people >67 years old in November 2015 in Sweden and focused on the 2 years prior to death. Women used LTC for 15.6 months and men for 14.1 months out of 24 these months. The length of stay in institutional care was 7.2 and 6.2 months, respectively. These survival data for LTC residents are in line with the estimated median survival of controls in our study, thus further validating the median survival in residents of LTC facilities is very limited.

We should acknowledge that there can be large heterogeneity in survival in different LTC facilities. Some LTC facilities admit mostly residents with known limited life-expectancy (mostly for palliative care), while others may be institutions that admit mostly older adults who are quite healthy or have limited health problems with substantial life-expectancy. A systematic review has found that across 6 cohort studies, the mortality rate within 6 months of admission to a nursing home ranged from 0% to 34% (median 20.2%).^19^ In our analysis, we could not include data on the features of each LTC facility (e.g. whether it focused on palliative care) and we could not match infected residents with uninfected residents from the same facility. Nevertheless, the control groups both in the main and in the sensitivity analysis seem to have median survival that is entirely compatible with the literature on LTC residents and their overall limited expected survival, on average.

Some additional caveats should be discussed. Our data pertain to fatalities during the first wave of COVID-19 and until the fall of 2020. The first wave was the most devastating in most high-income countries, with a few exceptions (e.g. Australia).^20,21^ The relatively lower proportion of fatalities in LTC residents in subsequent waves may reflect a combination of multiple factors: high levels of prior infection (seroprevalence studies have found 5-10 times higher infection rates in LTC facilities than in the general population in the first wave),^22-24^ better protection of nursing homes, more extensive testing, widespread use of vaccination in 2021,^21^ and the possibility that the sickest individuals were the first to succumb.^25^ Moreover, the lower risk of death after the first month post-infection versus the uninfected controls should not be interpreted as a sign that SARS-COV-2 infection causally decreases the risk of death during long-term follow-up, as it probably reflects mostly a selection process (residents who died in the first month were probably more sick and debilitated before infection, while those surviving probably had better life expectancy). Finally, we cannot exclude the possibility that some controls may have been asymptomatically infected but the infection remained unnoticed due to limited testing, especially in the early weeks of the pandemic. With more systematic testing after the end of the first wave and with limited epidemic activity during the late spring and summer of 2020, it is unlikely that infections in controls were missed in that period, let alone that these infections would shorten the survival of the control groups. As above, the observed limited survival of the control groups is entirely in line with data on residents of LTC facilities in the absence of COVID-19 from Sweden and elsewhere. In further support of our findings, excess death calculations for Sweden for 2020 and also for the entire pandemic period to end of 2021 and early 2022 show very limited excess deaths, if at all.^26,27^ This pattern is entirely congruent with the possibility that many/most residents who died of SARS-CoV-2 in the first wave had very limited life expectancy. Therefore, they would not contribute to excess death calculations, if excess deaths are assessed over 1-2 years downstream.

Allowing for these caveats, the major strength of our study is that it uses on large databases with nationwide coverage. Even so, similar analyses should also be performed in other countries because the health status of LTC residents may be different and with assessments covering also the vaccination period for a complete picture of the COVID-19 pandemic.^28^ This will allow to obtain more solid evidence on both the years-of-life-lost over 2020-2022, as well as insights about the long-term outcomes of SARS-CoV-2-infected residents of various types of LTC facilities who survived and recovered from the acute infection.

## Data Availability

The data files used for the present study is publicly unavailable according to regulations under Swedish law, but can be applied for at each of the respective registry holders.

## Author Contributions

Concept and design: MB, JPI, JB, PN, Acquisition, analysis, or interpretation of data: All authors. Drafting of the manuscript: MB, JPI. Critical revision of the manuscript for important intellectual content: All authors. Statistical analysis: MB and JB. Supervision: PN, JPI.

