## Supplemental Table 1 for "Time-Varying Risk of Death After SARS-CoV-2-Infection in Long-Term Care Facility Residents: A Matched Cohort Study"

| **Supplemental Table 1.** Baseline characteristics | | |  |  |
| --- | --- | --- | --- | --- |
| **Variables** | **SARS-CoV-2 infected residents (main analysis)** (n=3731) | **Uninfected  controls**  **(main analysis)**  (n=3731) | **SARS-CoV-2 infected residents (sensitivity)**  (n=3604) | **Uninfected controls (sensitivity)**  (n=3604) |
| **Median (IQR) days between Senior Alert registration and baseline^a^** | 120 (60-188) | 120 (60-188) | 118 (59-184) | 118 (59-184) |
| **Male sex** | 1325 (35.5) | 1318 (35.3) | 1278 (35.5) | 1233 (34.2) |
| **Age, median (IQR), yrs** | 87 (81-92) | 87 (81-92) | 86 (80-91) | 87 (81-92) |
| **Age group, yrs** |  |  |  |  |
| <70 | 140 (3.8) | 164 (4.4) | 152 (4.2) | 166 (4.6) |
| 70-74 | 249 (6.7) | 238 (6.4) | 251 (7.0) | 302 (8.4) |
| 75-79 | 456 (12.2) | 431 (11.6) | 465 (12.9) | 438 (12.2) |
| 80-84 | 706 (18.9) | 693 (18.6) | 965 (19.3) | 688 (19.1) |
| 85-89 | 938 (25.1) | 927 (24.9) | 911 (25.3) | 829 (23.0) |
| ≥90 | 1,242 (33.3) | 1,278 (34.3) | 1130 (31.4) | 1181 (32.8) |
| **BMI, kg/m^2^, mean (SD)** | 25.4 (5.0) | 25.6 (5.3) | 25.4 (5.1) | 25.0 (5.0) |
| **BMI categories** |  |  |  |  |
| Underweight (<18.5 kg/m^2^) | 240 (6.4) | 258 (6.9) | 233 (6.5) | 264 (7.3) |
| Normal weight (18.5-24.99 kg/m^2^) | 1,672 (44.8) | 1,604 (43.0) | 1614 (44.8) | 1701 (47.2) |
| Overweight (25.0-29.99 kg/m^2^) | 1,196 (32.1) | 1,182 (31.7) | 1108 (30.7) | 1160 (32.2) |
| Obesity (≥30 kg/m^2^) | 623 (16.7) | 687 (18.4) | 597 (14.7) | 531 (16.6) |
| **Neuropsychological conditions** |  |  |  |  |
| None | 886 (23.8) | 884 (23.7) | 858 (23.8) | 906 (25.1) |
| Mild dementia or depression | 1,822 (48.8) | 1,805 (48.4) | 1755 (48.7) | 1724 (47.8) |
| Severe dementia or depression | 1,023 (27.4) | 1,042 (27.9) | 991 (27.5) | 974 (27.0) |
| **Known previous falls** | 1,970 (52.8) | 1,950 (52.3) | 1893 (52.5) | 1884 (52.3) |
| **Walking ability** |  |  |  |  |
| Safe with or without walking aids | 1,513 (40.6) | 1,492 (40.0) | 1467 (40.7) | 1458 (40.5) |
| Unsafe walk | 1,367 (36.6) | 1,379 (37.0) | 1309 (36.3) | 1306 (36.2) |
| Unable to walk | 851 (22.8) | 860 (23.1) | 828 (23.0) | 840 (23.3) |
| **Fluid intake, ml/day** |  |  |  |  |
| >1000 | 2,191 (58.7) | 2,182 (58.5) | 2118 (58.8) | 2189 (60.7) |
| 700-1000 | 1,327 (35.6) | 1,312 (35.2) | 1292 (35.9) | 1237 (34.3) |
| 500-700 | 196 (5.3) | 210 (5.6) | 180 (5.0) | 161 (4.5) |
| <500 | 17 (0.5) | 27 (0.7) | 14 (0.4) | 17 (0.5) |
| **Food intake** |  |  |  |  |
| Normal serving | 2,597 (69.6) | 2.587 (69.3) | 2523 (70.0) | 2486 (69.0) |
| 3/4 serving | 686 (18.4) | 699 (18.7) | 654 (18.2) | 686 (19.9) |
| ½ serving | 350 (9.4) | 341 (9.1) | 334 (9.3) | 339 (9.4) |
| <½ serving | 98 (2.6) | 104 (2.8) | 93 (2.6) | 93 (2.6) |
| **General physical condition** |  |  |  |  |
| Good | 2,077 (55.7) | 2,034 (54.5) | 2020 (56.1) | 2002 (55.6) |
| Fair | 1,524 (40.9) | 1,545 (41.4) | 1463 (40.6) | 1471 (40.8) |
| Poor | 121 (3.2) | 142 (3.8) | 113 (3.1) | 128 (3.6) |
| Very bad | 9 (0.2) | 10 (0.3) | 8 (0.2) | 3 (0.1) |
| **Incontinence** |  |  |  |  |
| No | 986 (26.4) | 989 (26.5) | 952 (26.4) | 962 (26.7) |
| Temporarily but unusual | 565 (15.1) | 556 (14.9) | 542 (15.0) | 482 (13.4) |
| Urinary or bowel | 906 (24.3) | 890 (23.9) | 881 (24.5) | 940 (26.1) |
| Urinary and bowel | 1,274 (34.2) | 1,296 (34.7) | 1229 (34.1) | 1220 (33.9) |
| **Comorbidities** |  |  |  |  |
| Stroke | 942 (25.3) | 940 (25.2) | 911 (25.3) | 940 (26.1) |
| Myocardial infarction | 446 (12.0) | 431 (11.6) | 428 (11.9) | 408 (11.3) |
| Angina pectoris | 576 (15.4) | 574 (14.4) | 553 (15.3) | 550 (15.3) |
| Heart failure | 771 (20.7) | 776 (20.8) | 733 (20.3) | 721 (20.0) |
| Atrial fibrillation | 997 (26.7) | 971 (26.0) | 963 (26.7) | 936 (26.0) |
| Autoimmune disease | 487 (13.1) | 491 (13.2) | 454 (12.6) | 446 (12.4) |
| Diabetes | 825 (22.1) | 845 (22.7) | 791 (22.0) | 765 (21.2) |
| COPD | 483 (13.0) | 486 (13.0) | 454 (12.6) | 459 (12.7) |
| Asthma | 275 (7.4) | 242 (6.5) | 258 (7.2) | 247 (6.9) |
| Cancer | 1,687 (45.2) | 1,661 (44.5) | 1630 (45.2) | 1623 (45.0) |
| Renal failure/CKD | 521 (14.0) | 536 (14.4) | 479 (13.3) | 505 (14.0) |
| Liver disease | 72 (1.9) | 75 (2.0) | 65 (1.8) | 62 (1.7) |
| Sepsis | 316 (8.5) | 309 (8.3) | 298 (8.3) | 296 (8.2) |
| Influenza | 184 (4.9) | 193 (5.2) | 172 (4.8) | 174 (4.8) |
| Pneumonia | 915 (24.5) | 923 (24.7) | 870 (24.1) | 895 (24.8) |
| Alcohol intoxication | 233 (6.2) | 221 (5.9) | 226 (6.3) | 250 (6.9) |
| **Medications** |  |  |  |  |
| Antithrombotics | 2,205 (59.1) | 2,253 (60.4) | 2122 (58.9) | 2102 (58.3) |
| Antihypertensives (non-diuretic) | 2,257 (60.5) | 2,271 (60.9) | 2174 (60.3) | 2150 (59.7) |
| Diuretics | 1,611 (43.2) | 1,608 (43.1) | 1537 (42.7) | 1453 (40.3) |
| Antidepressants | 2,178 (58.4) | 2,140 (57.4) | 2100 (58.3) | 2078 (57.7) |
| Psycholeptics | 2,649 (71.0) | 2,648 (71.0) | 2556 (70.9) | 2553 (70.8) |
| The data are displayed as number (percent) unless stated otherwise. The data in the first two columns are the same as those presented in of our previous publication (reference 4).  Abbreviations: BMI, Body mass index; CKD, chronic kidney disease; COPD, chronic obstructive pulmonary disease; IQR, interquartile range; SD, standard deviation.  ^a^Baseline was the date of SARS-CoV-2 test/date of confirmed SARS-CoV-2 and the corresponding date in matched controls. | | | | |
